# The operational impact of deploying SARS-CoV-2 vaccines in countries of the WHO African Region

**DOI:** 10.1101/2020.08.13.20147595

**Authors:** Justin R. Ortiz, Joanie Robertson, Jui-Shan Hsu, Stephen L. Yu, Amanda J. Driscoll, Sarah R. Williams, Wilbur H. Chen, Meagan C. Fitzpatrick, Samba Sow, Robin J. Biellik, Jean-Marie Okwo-Bele, Kathleen M. Neuzil

## Abstract

**Background:** When available, SARS-CoV-2 vaccines will be deployed to countries with limited immunization systems.

**Methods:** We conducted an immunization capacity assessment of a simulated WHO African Region country using region-specific data on immunization, population, healthcare workers (HCWs), vaccine cold storage capacity (quartile values for national and subnational levels), and characteristics of influenza vaccines to represent future SARS-CoV-2 vaccines. We calculated monthly increases in vaccine doses, doses per vaccinator, and cold storage volumes for four-month SARS-CoV-2 vaccination campaigns targeting risk groups compared to routine immunization baselines.

**Findings:** Administering SARS-CoV-2 vaccines to risk groups would increase total monthly doses by 27.0% for ≥65 years, 91.7% for chronic diseases patients, and 1.1% for HCWs. Assuming median nurse density estimates adjusted for absenteeism and proportion providing immunization services, SARS-CoV-2 vaccination campaigns would increase total monthly doses per vaccinator by 29.3% for ≥65 years, 99.6% for chronic diseases patients, and 1.2% for HCWs. When we applied quartiles of actual African Region country vaccine storage capacity, routine immunization vaccine volumes exceeded national-level storage capacity for at least 75% of countries, but subnational levels had sufficient storage capacity for SARS-CoV-2 vaccines for at least 75% of countries.

**Interpretation:** In the WHO African Region, SARS-CoV-2 vaccination campaigns would substantially increase doses per vaccinator and cold chain capacity requirements over routine immunization baselines. Pandemic vaccination campaigns would add volume to national-level stores already at their limits, but sufficient capacity exists at subnational levels. Immediate attention to strengthening immunization systems is essential to support pandemic responses.

**Funding:** None

## BACKGROUND

There is a robust pipeline of SARS-CoV-2 vaccine candidates in various stages of preclinical and clinical development [1, 2]. While the challenges of developing a vaccine at an accelerated timeline for this newly-emerged pathogen have been well described [2], less attention has been given to the challenges of vaccine deployment and delivery. During the 2009 influenza A (H1N1) pandemic, vaccine deployment to many developing countries was suboptimal. In a survey about the 2009 H1N1 pandemic response published in 2013, WHO African Region (subsequently called “African Region”) countries reported 62% overall utilization of the 32,096,290 deployed H1N1 vaccine doses [3, 4], covering approximately 1% of the region’s population [4]. Most of these vaccine doses arrived after June 2010, despite the fact that H1N1 vaccines were prequalified by WHO for procurement beginning November 2009 [3]. This delay, largely due to obstacles in vaccine deployment and delivery, resulted in many preventable illnesses.

The SARS-CoV-2 pandemic has already caused more severe health impact than the 2009 H1N1 pandemic [5], and national governments, and technical and partner agencies aspire to higher population coverage than what was achieved during the prior pandemic [4, 6, 7]. A WHO report following the 2009 H1N1 pandemic concluded that existing health systems, immunization infrastructures, and deployment training played key roles during the vaccine response [8]. Critical to immunization infrastructure is the cold chain, defined as the series of actions and equipment necessary to maintain a vaccine within a specific low temperature range from production to the point of administration. While the number of deployed H1N1 vaccine doses covered only a fraction of at-risk populations identified, 29% of countries of the African Region reported having insufficient cold chain and logistics capacity for a comprehensive pandemic response [4].

Since 2014, a concerted global effort has improved cold chain infrastructure in developing countries [9, 10], which may facilitate a more rapid, widespread, and equitable SARS-CoV-2 vaccine response. We undertook this study to investigate the impact SARS-CoV-2 vaccination programs would have on vaccine delivery and storage systems in the African Region. Our goal was to assess the operational feasibility of mass vaccination programs targeting SARS-CoV-2 risk groups.

## METHODS

This analysis is similar to cold chain capacity assessments that are performed routinely by countries in preparation for new vaccine introductions [11]. We aimed to explore African Region-level operational concerns of SARS-CoV-2 vaccine implementation. We did not, however, intend for our analysis to take the place of country-level assessments using national contexts and data for deployment planning. Rather than describe any individual country or countries, we simulated a 20 million population country using the 2017 age distribution for the African Region [12]. We compared the number of doses and vaccine storage volumes required for routine immunization alone and with SARS-CoV-2 mass vaccination campaigns.

We then compared the required vaccine storage volumes for both scenarios to actual vaccine cold storage capacities from countries in the African Region. Our outcomes of interest included the monthly percentage increases in vaccine doses to be administered, doses administered per vaccinator, and cold storage volume requirements for SARS-CoV-2 mass vaccination campaigns compared to routine immunization baselines.

### Immunization strategies

We used a standard routine immunization schedule for the simulated country based on current WHO policy recommendations (Table 1) [13]. We then adopted a SARS-CoV-2 vaccination campaign strategy targeting the following risk groups: persons aged ≥65 years, persons with certain chronic medical conditions (chronic diseases), healthcare workers (HCWs), and all risk groups combined [7]. We assumed the SARS-CoV-2 vaccination campaign would span four months in duration and would require two doses, one month apart. To determine the fraction of the population in the chronic diseases group, we applied WHO-recommended [14], age-based estimates of comorbid risk factors for severe SARS-CoV-2 illness specific to the African Region [15]. Comorbidities included chronic cardiovascular, respiratory, neurologic, kidney and/or liver diseases; diabetes; cancers; HIV/AIDS; tuberculosis; and sickle cell disorders [15]. For HCWs, we used WHO estimates of skilled health personnel for the African Region, distributing these workers evenly across ages 20 through 64 years [16]. For the all risk groups combined category, we assumed HCWs had the same prevalence of chronic diseases as the general population and adjusted to avoid double counting.

**Table 1.** Routine and SARS-CoV-2 vaccines, schedules, and cold storage volume per dose

|  | Schedule | Cold storage tertiary packaging volume per dose (mL) | Cold storage secondary packaging volume per dose (mL) | Presentation | Product used |
| --- | --- | --- | --- | --- | --- |
| Routine infant immunization |  |  |  |  |  |
| Bacille Calmette-Guerin | Birth dose | 4.98 | 1.44 | 10 dose MDV | Japan BCG Laboratory |
| Hepatitis B | Birth dose | 12.56 | 2.86 | 10 dose MDV | Euvax B (paed) |
| Diphtheria-tetanus-pertussis-hepatitis B-Haemophilus influenzae type b (pentavalent) | 6, 10, and 14 weeks | 16.70 | 3.06 | 10 dose MDV | Eupenta |
| Polio (oral) | 6, 10, and 14 weeks | 6.22 | 1.40 | 10 dose MDV | PT Bio Farma (Persero) |
| Polio (inactivated) | 6 weeks | 23.95 | 4.00 | 5 dose MDV | Bilthoven Biologicals |
| Pneumococcal (conjugate) | 6, 10, and 14 weeks | 36.28 | 3.60 | 4 dose MDV | Pfizer |
| Rotavirus | 6 and 10 weeks | 49.45 | 46.30 | 1 dose SDV | Merck |
| Measles-rubella | 9-12 months, 13-24 months | 9.84 | 2.11 | 10 dose MDV | Serum Institute of India Pvt. Ltd. |
| Tetanus-diphtheria | 13-24 months | 9.47 | 2.38 | 10 dose MDV | PT Bio Farma (Persero) |
| Meningococcal A (conjugate) | 13-24 months | 9.84 | 2.11 | 10 dose MDV | Serum Institute of India Pvt. Ltd. |
| Yellow fever | 13-24 months | 3.59 | 2.99 | 10 dose MDV | Bio-Manguinhos/Fiocruz |
| Routine children immunization |  |  |  |  |  |
| HPV (girls only) | 2 doses from 9-14 years | 7.61 | 4.84 | 2 dose MDV | GlaxoSmithKline Biologicals SA |
| Tetanus-diphtheria | 9-14 years | 9.47 | 2.38 | 10 dose MDV | PT Bio Farma (Persero) |
| SARS-CoV-2 immunization targeting risk groups |  |  |  |  |  |
| ≥65 years | 2 doses, 1 month apart | 7.22 | 5.40 | 10 dose MDV | Seqirus Vaccines Limited |
| Healthcare workers | 2 doses, 1 month apart | 7.22 | 5.40 | 10 dose MDV | Seqirus Vaccines Limited |
| Chronic diseases | 2 doses, 1 month apart | 7.22 | 5.40 | 10 dose MDV | Seqirus Vaccines Limited |

**Notes:**
1. Vaccines and schedules are from WHO Immunization Tables except SARS-CoV-2 vaccines [37]. 2. Tertiary and secondary packaging volumes per dose from WHO Prequalified Vaccines Database with the exception of SARS-CoV-2 vaccines [17]. 3. SARS-CoV-2 cold storage volume per dose use prequalified 10-dose vial influenza vaccines as a proxy. 4. HPV vaccination programs generally target girls only aged 9 through 14 years [37]. For this simulation, we assumed the full HPV immunization series was given to girls 9 years of age.

### Vaccine storage

We identified WHO prequalified routine vaccine products [17], and we recorded the cold storage volumes required per dose for the standard routine immunization schedule (Table 1). We prioritized ten-dose vials for the schedule whenever they were available, choosing the product with the median secondary packaging volume if there were multiple prequalified ten-dose vial products. As no licensed SARS-CoV-2 vaccines are available, we adopted a proxy of cold storage volumes required for WHO prequalified influenza vaccines in ten-dose vials chosen similarly [17]. Cold storage volume is the total volume of vaccines maintained at refrigerated temperatures throughout storage and transport. Cold storage in countries is organized by levels, with the national level being where procured vaccines are received and stored before distribution to subnational levels (region, district, and health facility). Vaccines (and diluents, for reconstituted products) are produced by the manufacturer in vials or other primary packaging that are then packed together in labeled boxes called “secondary packaging” [17]. Products in secondary packaging are packed in cartons called “tertiary packaging” [17]. Vaccines are typically maintained in tertiary packaging at the national level [18]. Tertiary packaging volumes per dose are often around ten times the secondary packaging volumes per dose due to additional insulation and thermal packaging materials. At subnational levels, packaging materials are removed and vaccines are stored in their secondary packaging [18, 19]. Cold storage temperature for routine vaccines is typically 2° to 8°C, although some more thermostable products are becoming available [17]. Vaccine vials used in most developing countries carry individual thermo-chemical temperature monitors that indicate when a vial has been outside of the recommended storage temperature range and should be discarded [20, 21].

### Vaccine delivery

We estimated vaccine storage volumes using WHO tools and guidance [11, 17, 19, 20, 22]. We compared routine immunization to SARS-CoV-2 vaccination campaigns according to evidence-based assumptions about vaccine wastage (doses that are damaged or unused) [23], reserve stock (excess supply in case of increased demand or stock outs) [11, 20, 24], and resupply intervals (Table 2) [22]. In the case of SARS-CoV-2 vaccines, we assumed 7.5% wastage, 0% reserve stock, and monthly resupply intervals given anticipated high global demand and limited supply. We received summary vaccine cold storage capacity data from Gavi, the Vaccine Alliance, for African Region countries that are eligible to receive Gavi support. These data included the minimum, 25^th^ percentile, 50^th^ percentile, 75^th^ percentile, and upper range values of total country storage capacities, as well as the median proportion of national-level stores and health facility-level stores to total country capacity. For comparisons, capacity data were standardized by dividing the total cold storage capacity by the population of children aged < 2 years in each country.

**Table 2.** Study assumptions

|  | **Routine immunization assumption** | **SARS-CoV-2 immunization assumption** | **Comments** |
| --- | --- | --- | --- |
| **Vaccines** | WHO prequalified vaccines in multidose vial presentation given per WHO recommended schedule.<sup>14</sup> | SARS-CoV-2 vaccine in multidose vial presentation given as two doses, one month apart. | Given that there are no licensed SARS-CoV-2 vaccines, dose assumptions rely on dose schedule of SARS-CoV-2 vaccine clinical trials in the United States to date, and the volume analyses use packaging volumes per dose of WHO prequalified influenza vaccines as a proxy. |
| **Packaging** | Tertiary packaging at the national level<br>Secondary packaging at all subnational levels | Tertiary packaging at national level<br>Secondary packaging at all subnational levels | National level vaccine volume analyses use total tertiary packaging volume required per dose (the unit for international transport), defined as the volume of the container holding cartons which contain vaccine vials divided by the total doses contained.<br>Subnational levels vaccine volume analysis use total secondary packaging volume required per dose, defined as the volume of cartons which contain vaccine vials divided by the total doses contained. |
| **Storage temperature** | 2° to 8°C | 2° to 8°C |  |
| **Coverage** | 90% of target group | 90% of target group |  |
| **Target groups** | WHO recommended ages | Chronic diseases (any age)<br>Persons ≥ 65 years<br>Healthcare workers | Adults with chronic disease and older adults are at increased risk for severe SARS-CoV-2 disease [7]. Health care workers are at increased risk for SARS-CoV-2 infection and disease [7]. |
| **Strategy** | Year round | Four month mass vaccination campaign | Once pandemic vaccines are available, there will be an imperative to deliver them expeditiously. |
| **Wastage multidose vials** | 25% | 7.5% | Vaccine wastage is the doses that are lost or unused. Routine immunization inputs are from WHO guidance while SARS-CoV-2 inputs are from WHO Vaccine Wastage Rates Calculator assuming African Region, four weeks of retention of vaccine after vial opened, 2 dose schedule, 90% coverage, daily immunization sessions, and influenza vaccine [23]. |
| **Reserve stock** | 3 months at national level<br>1 month at district and regional levels<br>0.5 months at health facility level | No reserve stock | Vaccine reserve stock are the excess supply in case of increased demand or stock-outs. Routine inputs are from WHO guidance while SARS-CoV-2 inputs assume high global demand and limited supply [11, 20, 24]. |
| **Resupply intervals** | 3 months at national, district, and regional levels<br>1 month at health facility level | Every month | Three month supply interval is common for routine immunization in low resource settings [22], while the SARS-CoV-2 supply interval assumes high global demand and limited supply. |
| **Vaccinators** | Nurse density per capita for WHO African Region countries multiplied by the simulated country population, the proportion of nurses providing immunization services, and estimates of absenteeism at baseline and during the SARS-CoV-2 pandemic | Same as for routine | Typically, persons delivering vaccines in the region are nurses, but not all nurses engage in the provision of immunization services. The estimates for nurses per capita are from WHO [16]. Estimates of percentage of nurses providing immunization services are from the Organisation for Economic Co-operation and Development [25]. Absenteeism estimates from observational data (baseline) and modelling data (SARS-CoV-2) from the United States [26, 27]. |

### Analysis

We calculated the monthly doses and cold storage volumes required for routine immunization programs and SARS-CoV-2 vaccination campaigns to reach 90% of target populations [19]. We developed a vaccine flow-down schematic (Supplemental Figure 1) to depict the quantity of routine vaccine doses maintained at each immunization system level by month under normal circumstances and used it to calculate the total quantity of vaccine doses and their storage volumes at each level. To assess the feasibility of storing SARS-CoV-2 vaccines, we compared the anticipated vaccine volumes for routine immunization and SARS-CoV-2 vaccination campaigns to actual vaccine cold storage capacities at national and subnational levels under normal circumstances. We defined “immunization program workloads” as the doses delivered per vaccinator. To determine this, we calculated the number of monthly doses for each activity divided by the estimated number of vaccinators available in the country. We obtained estimates of the number of nurses for countries from a WHO Workforce Database [16], and we calculated nurse density per 10,000 population using 2017 country population estimates [12]. We determined the values for 25^th^ percentile, 50^th^ percentile, 75^th^ percentile, and upper range nurse density for African Region countries, and we determined median values for other WHO Regions. To assess the range of immunization program workloads that would be experienced across the African Region, we applied the different nurse density estimates to the simulated country. We multiplied each nurse density estimate by 46%, the proportion of nurses estimated to provide immunization services [16, 25]. We assumed a 3% baseline absenteeism and an additional 8% absenteeism for SARS-CoV-2 pandemic months [26, 27]. To place results within the global context, we compared these results to calculations using the median number of vaccinators per capita from other WHO regions, while maintaining all other parameters unchanged. For the monthly comparisons and time series, we assumed constant routine immunization demand throughout the year and constant SARS-CoV-2 immunization demand during a mass vaccination campaign of four months duration. We performed all analyses using standard spreadsheet software including the made-for-purpose “WHO Vaccine Volume Calculator 2012” (Microsoft Corp, Redmond, WA, US) [28]. The WHO Vaccine Volume Calculator is designed to estimate the net storage volume of vaccines per child. Its specific purpose is to analyze how changes to the national immunization program schedule will affect cold chain capacity. As there was no involvement of human participants or personal identifiable information, institutional review board approval was not required.

**Figure 1.**
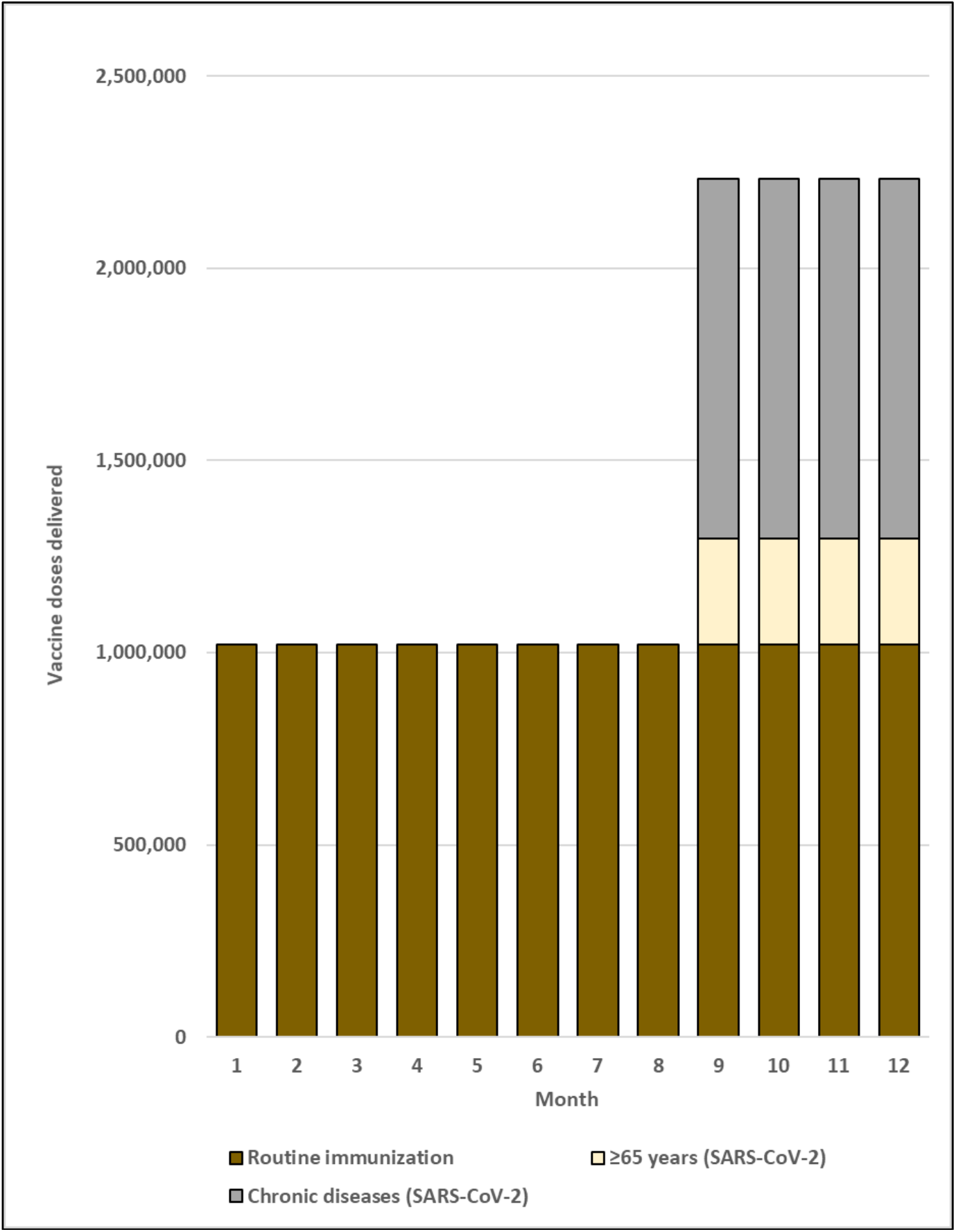
Monthly doses delivered for routine and SARS-CoV-2 risk group vaccination. Notes: 1. Figure includes sufficient doses for 90% coverage of target groups. 2. Figure assumes constant routine immunization demand over the year and constant SARS-CoV-2 immunization demand over a four-month mass vaccination campaign. 3. For illustrative purposes only, the mass vaccination campaign is conducted during months 9 through 12. 4. HCWs target group removed because volume contribution was too small to depict in the Figure.

## RESULTS

### Description of population and vaccination target groups

We simulated a country of 20 million people in the African Region (Table 3). The percentage of the total population within each risk group was 3.1% for persons ≥65 years, 10.4% for persons with chronic diseases, 0.1% for HCWs, and 11.8% for all risk groups combined.

**Table 3.** Total and SARS-CoV-2 vaccine target group populations by age group

| Age group | Total population | ≥65 years |  | Chronic diseases |  | HCWs |  | All risk groups combined |  |
| --- | --- | --- | --- | --- | --- | --- | --- | --- | --- |
|  |  | n | % of total age group population | n | % of total age group population | n | % of total age group population | n | % of total age group population |
| Under 5 | 3,139,586 | 0 | 0.0% | 50,940 | 1.6% | 0 | 0.0% | 50,940 | 1.6% |
| 5 to 9 | 2,828,076 | 0 | 0.0% | 45,886 | 1.6% | 0 | 0.0% | 45,886 | 1.6% |
| 10 to 14 | 2,500,943 | 0 | 0.0% | 40,578 | 1.6% | 0 | 0.0% | 40,578 | 1.6% |
| 15 to 19 | 2,128,524 | 0 | 0.0% | 240,909 | 11.3% | 0 | 0.0% | 240,909 | 11.3% |
| 20 to 24 | 1,817,105 | 0 | 0.0% | 205,662 | 11.3% | 2,842 | 0.2% | 208,183 | 11.5% |
| 25 to 29 | 1,559,398 | 0 | 0.0% | 176,495 | 11.3% | 2,842 | 0.2% | 179,015 | 11.5% |
| 30 to 34 | 1,331,654 | 0 | 0.0% | 150,718 | 11.3% | 2,842 | 0.2% | 153,239 | 11.5% |
| 35 to 39 | 1,109,669 | 0 | 0.0% | 125,594 | 11.3% | 2,842 | 0.3% | 128,114 | 11.5% |
| 40 to 44 | 898,313 | 0 | 0.0% | 101,672 | 11.3% | 2,842 | 0.3% | 104,193 | 11.6% |
| 45 to 49 | 717,631 | 0 | 0.0% | 81,222 | 11.3% | 2,842 | 0.4% | 83,743 | 11.7% |
| 50 to 54 | 571,040 | 0 | 0.0% | 174,724 | 30.6% | 2,842 | 0.5% | 176,697 | 30.9% |
| 55 to 59 | 448,210 | 0 | 0.0% | 169,635 | 37.8% | 2,842 | 0.6% | 171,402 | 38.2% |
| 60 to 64 | 338,046 | 0 | 0.0% | 156,064 | 46.2% | 2,842 | 0.8% | 157,595 | 46.6% |
| 65 to 69 | 238,867 | 238,867 | 100.0% | 134,012 | 56.1% | 0 | 0.0% | 238,867 | 100.0% |
| 70+ years | 372,940 | 372,940 | 100.0% | 225,615 | 60.5% | 0 | 0.0% | 372,940 | 100.0% |
| Total | 20,000,000 | 611,807 | 3.1% | 2,079,729 | 10.4% | 25,580 | 0.1% | 2,352,301 | 11.8% |

Notes:
1. Total population of simulated country with the age distribution of the African Region per 2017 [12]. 2. From estimates of chronic diseases risk factors for severe SARS-CoV-2 illness for Africa [15]. 3. HCWs population used per capita estimates from the WHO African Region (12.79 per 10,000 population) multiplied by the simulated country population and distributed evenly from ages 20 to 64 [16]. 4. The sum of populations in the ≥65 years, chronic diseases, and HCW groups does not equal the population of all risk groups combined, as the HCW population was adjusted to account for persons with chronic diseases.

### Vaccine doses

As in most developing countries, all routine vaccination services in the simulated country would be received by children aged <5 years and 9 through 14 years. Assuming systems would also be in place to provide immunization services to HCWs, established routine immunization platforms would reach 31.0% of the national population. A total of 1,020,931 routine vaccine doses would be given each month. Each month during the SARS-CoV-2 vaccination campaign, 275,313 doses would be delivered targeting persons ≥65 years, 935,878 targeting persons with chronic diseases, 11,511 targeting HCWs, and 1,058,536 targeting all risk groups combined (Table 4 and Figure 1). Targeting SARS-CoV-2 vaccination of risk groups would increase monthly doses administered over the routine immunization baseline by 27.0% for persons ≥65 years, 91.7% for persons with chronic diseases, 1.1% for HCWs, and 103.7% for all risk groups combined.

**Table 4.** Monthly vaccine doses for routine immunization and SARS-CoV-2 mass vaccination campaigns by target group

| Age group | Routine immunization (doses) | ≥65 years (SARS-CoV-2 vaccine doses) | Chronic diseases (SARS-CoV-2 vaccine doses) | HCWs (SARS-CoV-2 vaccine doses) | All risk groups combined (SARS-CoV-2 vaccine doses) |
| --- | --- | --- | --- | --- | --- |
| Under 5 | 939,468 | 0 | 22,923 | 0 | 22,923 |
| 5 to 9 | 81,462 | 0 | 20,649 | 0 | 20,649 |
| 10 to 14 | 0 | 0 | 18,260 | 0 | 18,260 |
| 15 to 19 | 0 | 0 | 108,409 | 0 | 108,409 |
| 20 to 24 | 0 | 0 | 92,548 | 1,279 | 93,682 |
| 25 to 29 | 0 | 0 | 79,423 | 1,279 | 80,557 |
| 30 to 34 | 0 | 0 | 67,823 | 1,279 | 68,958 |
| 35 to 39 | 0 | 0 | 56,517 | 1,279 | 57,651 |
| 40 to 44 | 0 | 0 | 45,753 | 1,279 | 46,887 |
| 45 to 49 | 0 | 0 | 36,550 | 1,279 | 37,684 |
| 50 to 54 | 0 | 0 | 78,626 | 1,279 | 79,514 |
| 55 to 59 | 0 | 0 | 76,336 | 1,279 | 77,131 |
| 60 to 64 | 0 | 0 | 70,229 | 1,279 | 70,918 |
| 65 to 69 | 0 | 107,490 | 60,305 | 0 | 107,490 |
| 70+ years | 0 | 167,823 | 101,527 | 0 | 167,823 |
| Total | 1,020,931 | 275,313 | 935,878 | 11,511 | 1,058,536 |
| Immunization program workload | Routine immunization (monthly doses / vaccinator) | ≥65 years (monthly SARS-CoV-2 vaccine doses / vaccinator) | Chronic diseases (monthly SARS-CoV-2 vaccine doses / vaccinator) | HCWs (monthly SARS-CoV-2 vaccine doses / vaccinator) | All risk groups combined (monthly SARS-CoV-2 vaccine doses / vaccinator) |
| Routine immunization (prepandemic baseline) | 165.3 | -- | -- | -- | -- |
| Routine immunization (pandemic) | 179.7 | 48.5 | 164.7 | 2.0 | 186.3 |
| % of routine immunization prepandemic baseline | 108.7% | 29.3% | 99.6% | 1.2% | 112.7% |

Notes:
1. Analysis assumes 90% coverage of target groups and a two-dose series for SARS-CoV-2 vaccines 2. We assumed that both HPV doses were delivered to girls at 9 years of age. 3. Vaccinators = nurse density per capita (6.9 per 10,000) x country population (20,000,000) x percent of nurses that provide immunization services (46%). Nurse data are from WHO Global Health Workforce Statistics [16]. We used the median value from African Region countries at the most recent available date and divided by country population estimates to calculate per capita values [12]. 4. Adjustments account for 3% absenteeism (baseline) and an additional 8% absenteeism (during pandemic) [26, 27].

### Immunization workload

Using the median value of nurse density from countries in the African Region (6.9 per 10,000 population), we estimated 13,840 nurses in the simulated country, of which an estimated 6,366 (46%) would provide vaccination services. Accounting for baseline and pandemic absenteeism, one vaccinator would administer 165.3 routine doses per month at baseline and 179.7 routine doses per month during the pandemic (Table 4).

During the SARS-CoV-2 vaccination campaign, the number of persons vaccinated monthly per vaccinator would be 48.5 targeting persons ≥65 years, 164.7 targeting persons with chronic diseases, 2.0 targeting HCWs, and 186.3 targeting all risk groups combined. Compared to routine vaccines given at baseline, SARS-CoV-2 vaccination of risk groups would increase the immunization program workload for vaccinators by 29.3% for persons ≥65 years, 99.6% for persons with chronic diseases, 1.2% for HCWs, and 112.7% for all risk groups combined. Due to nursing personnel shortages in the African Region, the immunization program workload would be substantially lower if nurse density were similar to other WHO Regions. Repeating the analysis using nurse density estimates from other WHO Regions (but maintaining all other parameters unchanged), the immunization program workload would increase by 26.9% using estimates from the Americas, 36.6% using estimates from the Eastern Mediterranean, 10.4% using estimates from Europe, 42.2% using estimates from South-East Asia, 19.8% using estimates from the Western Pacific, and 24.3% using global estimates (with the relative proportion changes being the same regardless of risk group) (Supplemental Table 2 and Figure 2).

**Figure 2.**
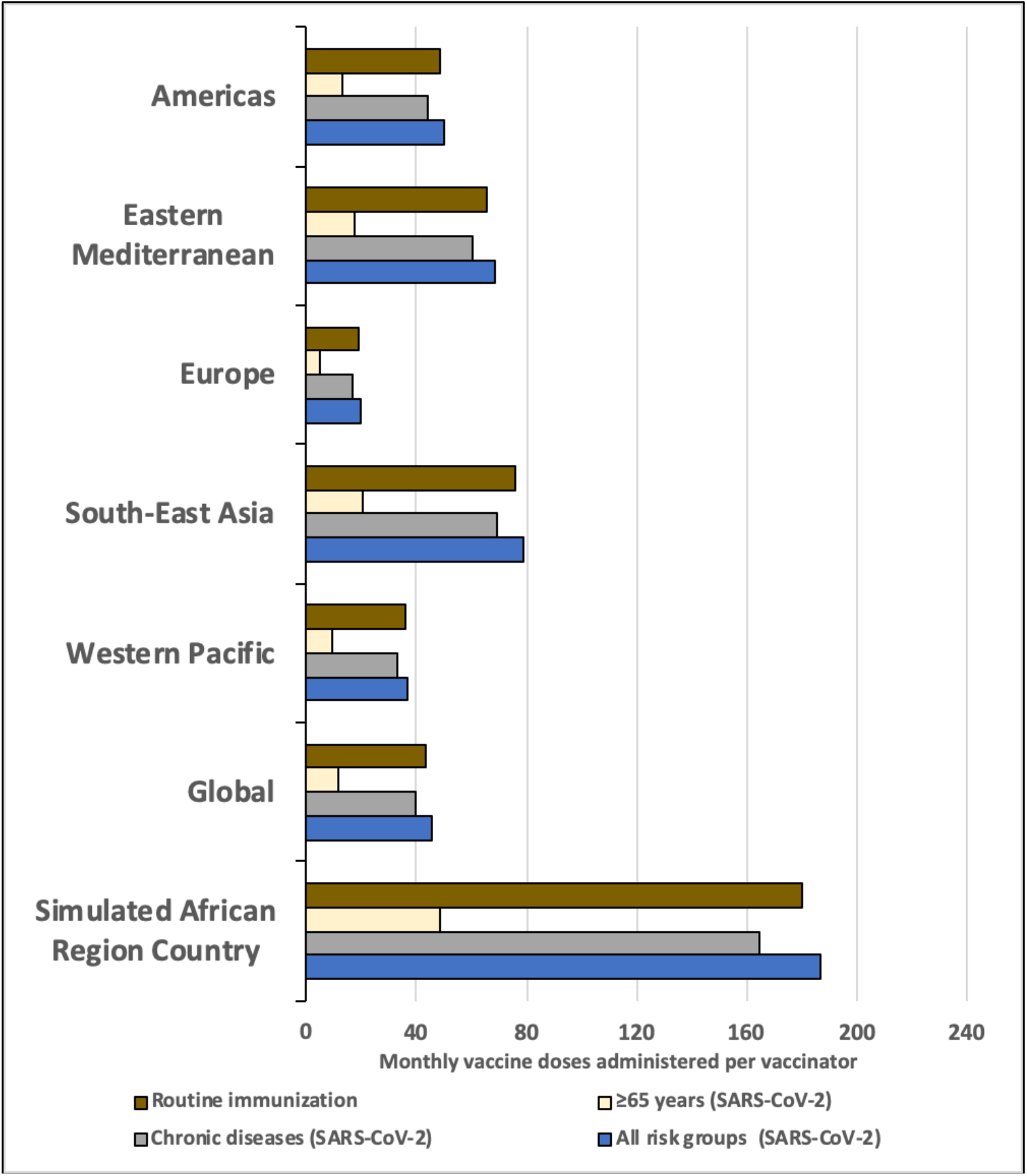
Monthly routine and SARS-CoV-2 campaign vaccine doses per vaccinator, by WHO Region estimated nurse density. Notes: 1. Data points represent the total monthly vaccines delivered divided by the total number of vaccinators in a country. Vaccinators = nurse density per capita x country population (20,000,000) x percent of nurses that provide immunization services (46%). 2. Nurse data are from WHO Global Health Workforce Statistics [16]. We used the median value from countries at the most recent available date and divided by country population estimates [12] to calculate per capita values and then categorized by WHO Region. 3.Routine immunization programs account for 3% absenteeism (baseline). SARS-CoV-2 vaccination programs account for 3% absenteeism (baseline) and an additional 8% absenteeism (during pandemic). 4. HCWs target group removed because volume contribution was too small to depict in the figure.

We repeated the workload analysis using quartiles and upper range estimates for nurse density from countries in the African Region (while maintaining all other parameters unchanged). For each nurse density value, we estimated the following number of vaccinators: 25^th^ percentile: 4,177; 50^th^ percentile: 6,366; 75^th^ percentile: 12,654; and upper range value: 62,957 (Supplemental Table 3 and Supplemental Figure 4). The 75^th^ percentile nurse density estimate (13.8 per 10,000 population) was still lower than all other WHO Region nurse density estimates (with the closest being South-East Asia at 16.4 per 10,000 population). The upper range value (68.4 per 10,000 population) exceeded the nurse density estimate for all other WHO Regions (i.e. higher than the 66.5 per 10,000 population estimate for the European Region). Additional monthly doses per vaccinator for SARS-CoV-2 risk groups ranged from 4.9-73.9 targeting persons ≥65 years, 16.7-251.1 targeting persons with chronic diseases, 0.2-3.1 targeting HCWs, and 18.8-284.0 targeting all risk groups combined.

**Figure 3.**
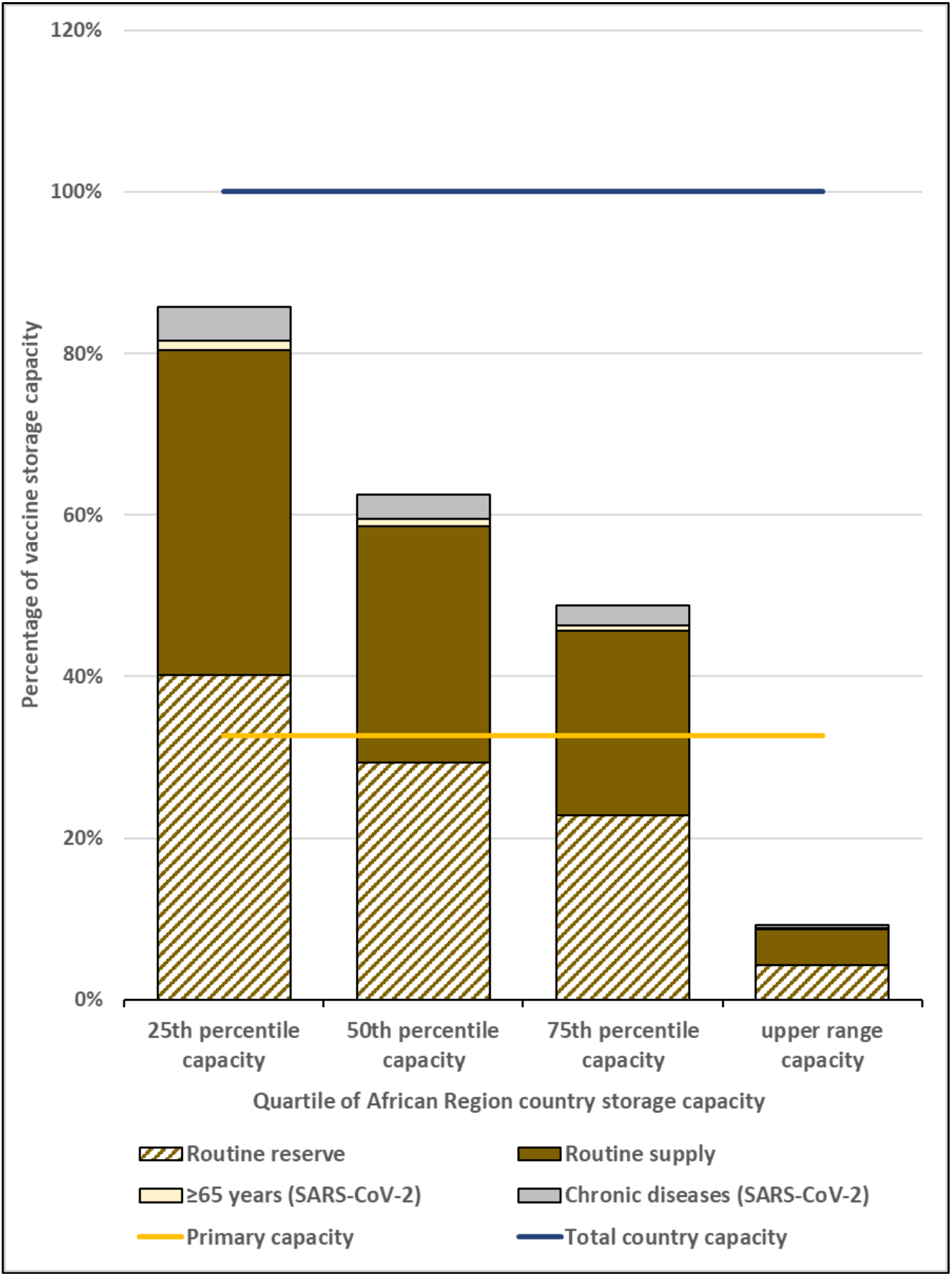
Maximum national level monthly routine and SARS-CoV-2 vaccine volumes as a proportion of total country capacity, by quartile of African Region country storage capacity. Notes: 1. Based on the maximum monthly national level vaccine storage volume (month 10) of the vaccine flow-down schematic (Figure 1). 2. HCWs target group removed because volume contribution was too small to depict in the figure.

**Figure 4.**
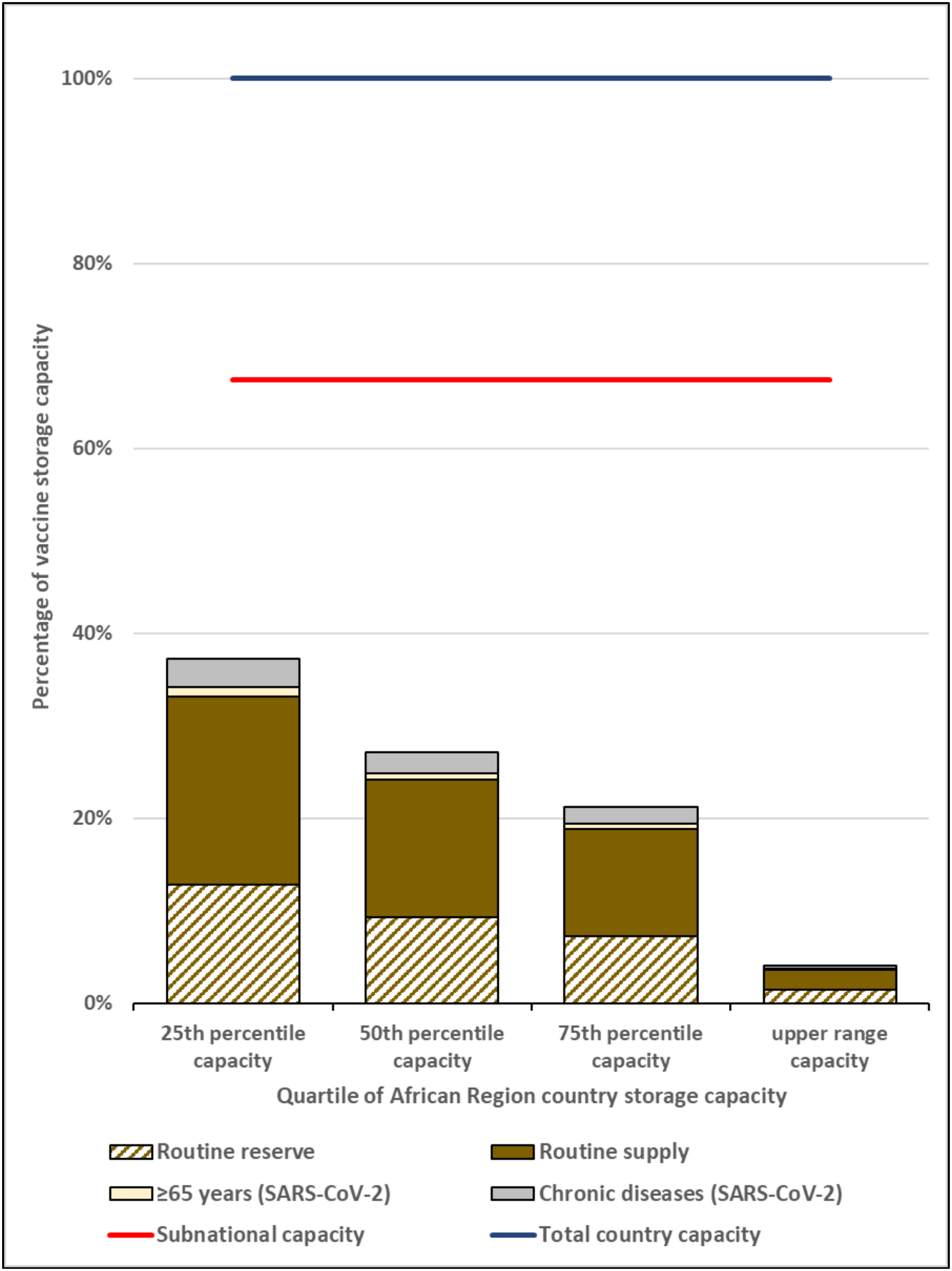
Maximum subnational level monthly routine and SARS-CoV-2 vaccine volumes as a proportion of total country capacity, by quartile of African Region country storage capacity. Notes: 1. Based on the maximum monthly subnational vaccine storage volume (month 11) of the vaccine flow-down schematic (Figure 1). 2. HCWs target group removed because volume contribution was too small to depict in the figure.

### Routine vaccine storage

We applied standardized African Region vaccine cold storage capacity range and quartiles to the simulated country. The total storage capacity would range from 38,403 L to 1,605,826 L, and national-level storage capacity would range from 12,535 L to 524,141 L (Table 5). We assessed the storage capacity during the month with the highest vaccine volumes. Using the vaccine flow-down schematic and adding the SARS-CoV-2 vaccination campaign during months 9 through 12, the maximum vaccine volumes were during month 10 for the national level and month 11 for the subnational levels (Supplemental Figures 2 and 3). We found that the maximum monthly routine vaccine volumes for the national level would be 139,161 L and for the aggregate subnational levels would be 57,472 L. At the national level, only the highest value capacity estimate had sufficient cold storage capacity to accommodate all routine vaccines according to our vaccine storage assumptions. Routine vaccine volumes exceeded that available at the national level for at least 75% of African Region country storage capacities (Figure 3). At subnational levels, substantial excess available space existed for all capacities assessed, indicating that subnational levels had sufficient storage capacity for SARS-CoV-2 vaccines for at least 75% of countries (Figure 4).

**Table 5.** Maximum monthly storage volume for routine and SARS-CoV-2 vaccines by quartile of African Region country storage capacity

|  |  | **Lower range capacity** | **25th percentile capacity** | **50th percentile capacity** | **75th percentile capacity** | **Upper range capacity** |
| --- | --- | --- | --- | --- | --- | --- |
| **Country vaccine storage capacity** | % of total country capacity | volume (L) | volume (L) | volume (L) | volume (L) | volume (L) |
| Total country capacity | 100.0% | 38,403 | 173,159 | 237,433 | 304,609 | 1,605,826 |
| National level | 32.6% | 12,535 | 56,519 | 77,498 | 99,424 | 524,142 |
| Subnational level | 67.4% | 25,868 | 116,640 | 159,935 | 205,185 | 1,081,685 |
| **National level maximum monthly vaccine volumes** | Vaccine volume (L) | % of national level capacity | % of national level capacity | % of national level capacity | % of national level capacity | % of national level capacity |
| Routine immunization | 139,161 | 1110.2% | 246.2% | 179.6% | 140.0% | 26.6% |
| ≥65 years (SARS-CoV-2) | 2,137 | 17.0% | 3.8% | 2.8% | 2.1% | 0.4% |
| Chronic diseases (SARS-CoV-2) | 7,264 | 57.9% | 12.9% | 9.4% | 7.3% | 1.4% |
| HCWs (SARS-CoV-2) | 89 | 0.7% | 0.2% | 0.1% | 0.1% | <0.1% |
| All risk groups combined (SARS-CoV-2) | 8,216 | 65.5% | 14.5% | 10.6% | 8.3% | 1.6% |
| **Subnational level maximum monthly vaccine volumes** | Vaccine volume (L) | % of subnational level capacity | % of subnational level capacity | % of subnational level capacity | % of subnational level capacity | % of subnational level capacity |
| Routine immunization | 57,472 | 222.2% | 49.3% | 35.9% | 28.0% | 5.3% |
| ≥65 years (SARS-CoV-2) | 1,598 | 6.2% | 1.4% | 1.0% | 0.8% | 0.1% |
| Chronic diseases (SARS-CoV-2) | 5,433 | 21.0% | 4.7% | 3.4% | 2.6% | 0.5% |
| HCWs (SARS-CoV-2) | 67 | 0.3% | 0.1% | 0.0% | 0.0% | <0.1% |
| All risk groups combined (SARS-CoV-2) | 6,145 | 23.8% | 5.3% | 3.8% | 3.0% | 0.6% |

Notes:

### SARS-CoV-2 vaccine storage

We estimated the volume required to store SARS-CoV-2 vaccines in tertiary packaging at the national level and in secondary packaging at subnational levels (Supplemental Figures 2 and 3). At the national level, monthly SARS-CoV-2 vaccines would occupy 2,137 L for persons ≥65 years, 7,264 L for persons with chronic diseases, 89 L for HCWs, and 8,216 L for all risk groups combined (Table 5). Excluding the lowest range value for country capacity, the percentage of total national-level stores displaced by these vaccines by quartile of African Region country capacities ranged from 0.4% to 3.8% for persons ≥65 years, 1.4% to 12.9% for persons with chronic diseases, <0.1% to 0.2% for HCWs, and 1.6% to 14.5% for all risk groups combined (Table 5 and Figure 3). At the subnational levels, monthly SARS-CoV-2 vaccines targeting risk groups would occupy 1,598 L for persons ≥65 years, 5,433 L for persons with chronic diseases, 67 L for HCWs, and 6,145 L for all risk groups combined. The percentage of subnational-level stores displaced by these vaccines by quartile of African Region country storage capacity ranged from 0.1% to 6.2% for persons ≥65 years, 0.5% to 21.0% for persons with chronic diseases, <0.1% to 0.3% for HCWs, and 0.6% to 23.8% for all risk groups combined (Table 5 and Figure 4).

## DISCUSSION

We used WHO tools and guidelines to estimate the operational impact of a SARS-CoV-2 vaccination campaign in the African Region [17, 22]. While we simulated a country of 20 million population for this analysis, we used actual immunization, population, healthcare worker density, and cold storage capacity data available for the region. As no SARS-CoV-2 vaccines are currently licensed [1], we assumed a two dose vaccine series, multidose vials, and vaccine storage and volume characteristics similar to influenza vaccines.

Our analysis revealed that the national-level storage capacity would be insufficient to accommodate additional SARS-CoV-2 vaccines for a vaccination campaign targeting risk groups as large as persons ≥65 years (3.1% of the population) in at least 75% of African Region countries. However, all African Region countries likely have sufficient subnational-levels storage capacity to accommodate SARS-CoV-2 vaccines for mass vaccination campaigns. This means that there is insufficient excess cold storage capacity at the national level, whereas once vaccines are distributed to subnational levels in the health system there should be adequate storage capacity. This bottleneck in the vaccine cold chain at the national level can be addressed in advance by decreasing reserve stocks, shortening supply intervals, removing products from tertiary packaging earlier in the chain, distributing vaccines to the subnational levels, and/or installing or leasing more refrigeration capacity. However, implementation of SARS-CoV-2 vaccination campaigns will also require increases in accompanying dry goods, such as syringes, safety boxes, cotton, alcohol, and personal protective equipment, necessitating additional planning and budget for operations and waste management. More in-country vaccine shipments will be required to distribute routine and SARS-CoV-2 vaccines if transportation capacities are not increased, adding costs and logistical challenges. Care should be taken not to disrupt routine vaccine supply and distribution, as stockouts of routine vaccines will exacerbate national immunization programs already impacted by the pandemic.

While we found that the aggregate subnational-level storage capacities are adequate for SARS-CoV-2 vaccines, with very large margins expected for a majority of countries, it should be noted that it is not possible nor practical to fill all available subnational-level storage capacity. For example, at the health facility level, standard vaccine refrigerators may have multiple times the capacity needed for vaccines used at that facility, but that excess capacity cannot be easily used to store vaccines for other facilities.

The complicated logistics of fully utilizing the available storage capacity at the lower levels of the cold chain makes it impractical. Nevertheless, our findings show that there is ample vaccine storage capacity available at subnational levels, and creative management of both routine and campaign vaccines could leverage some of that excess capacity during the SARS-CoV-2 vaccination campaign period.

Another major obstacle for SARS-CoV-2 vaccine delivery in the African Region will be the availability of qualified vaccinators. The African Region has the smallest health workforce of any WHO Region [16], and the workload required to vaccinate risk groups would increase by 27.0% for persons ≥65 years and 91.7% for persons with chronic diseases, compared to routine immunization baselines. SARS-CoV-2 vaccination activities targeting HCWs will require less additional workload given their small population size and ease of access to vaccination services. Our analysis highlights the need for significant planning, and investment in equipment, training, and logistics for SARS-CoV-2 vaccine deployment.

There is reason for optimism that the African Region SARS-CoV-2 vaccination response will be much improved from the 2009 H1N1 pandemic experience. Over the last ten years, a concerted global effort has greatly improved cold chain infrastructures and increased storage capacity [9, 10]. Furthermore, countries in the African Region have successfully implemented mass vaccination campaigns with several different vaccines. From 2010-2017, the MenAfriVac introduction strategy targeted all persons aged one through 29 years in 23 Meningitis Belt countries [29]. From 2010-2011, six West African countries implemented mass vaccination campaigns achieving 98% coverage in the target population [29]. Over a ten-day period in Burkina Faso, a team of 5,328 vaccinators and as many volunteers vaccinated 11,425,391 persons (73% of the country population) [29]. The lyophilized vaccine was supplied in 10-dose vials, requiring reconstitution [17]. Each vaccinator vaccinated an average of 214 persons daily. The Region has had many other successful vaccination campaigns in the last decade, as well, including with vaccination against polio, measles, and yellow fever [30-33]. The impact on cold storage and workforce is also likely to be reduced by SARS-CoV-2 vaccine supply limitations, as WHO is currently planning for an initial tranche of vaccines to cover 3% of a country’s population [34]. With sufficient planning and resources, success could be achieved for SARS-CoV-2 vaccination programs targeting much higher proportion of a country’s population.

A SARS-CoV-2 vaccination campaign would be greatly facilitated if vaccine delivery were simplified [21]. While a one dose vaccine would be ideal, initial Phase 1 data indicate that two doses will likely be needed [2]. Nasal spray or transdermal patch vaccines that could be administered by trained lay persons would free skilled health workers to provide other essential services, but no such vaccines are currently in human trials [1]. While single dose vial vaccine presentations limit vaccine wastage [4, 20], and prefilled syringe presentations limit wastage and greatly simplify administration [20], their cold storage volumes per dose make these presentations highly problematic for use in mass vaccination campaigns in the African Region. For influenza vaccines (our proxy for SARS-CoV-2 vaccines in this analysis), secondary and tertiary packaging volumes per dose for multidose vials (5.4 mL and 7.2 mL) [17] are much lower than for single dose vials (median of WHO prequalified products: 18.4 mL and 87.3 mL) [17] and prefilled syringes (Flublok® Quadrivalent: 86.5 mL and 111.8 mL) (personal communication Global Medical Information, Sanofi Pasteur Inc).

WHO has advised that certain products would be challenging to include in national immunization schedules if they do not meet specific Programmatic Suitability Criteria [35]. Vaccine products must meet these mandatory criteria to achieve WHO prequalification: include an anti-microbial preservative (for injectable liquid vaccines in multi-dose containers), be relatively thermostable (not requiring storage at less than −20°C), have a dose volume ≤1 ml (for injectable vaccines), and not require intravenous administration [35]. Exceptions to the programmatic suitability criteria are made for the prequalification of certain emergency-use vaccines, such as for the prevention of Ebola Virus Disease, which are stored at −80°C to −60°C [29]. The first SARS-CoV-2 vaccine candidates to enter human trials in the United States used a mRNA platform [1, 2]. Most mRNA vaccines for early phase studies are frozen at −70°C [36], although more stable product are anticipated to be used for the later phase trials. The final storage temperature for any future licensed mRNA vaccines is unknown. DNA vaccine candidates are more thermostable, but would likely require administration via electroporation devices. Any licensed products requiring freezing for storage or nonstandard devices for vaccine administration would be prohibitive to use at scale in developing countries.

Our study should be interpreted in light of its strengths and limitations. Our use of real-world data inputs and WHO tools and guidance strengthen the generalizability of the results and comparability to other cold chain capacity assessments. Since there is no currently licensed SARS-CoV-2 vaccine, we used the characteristics of influenza vaccines as proxies. If a SARS-CoV-2 vaccine requiring only one dose for protection is eventually licensed, the estimated doses and storage volumes required could substantially decrease. Conversely, if only products requiring ultra-low temperature cold chain or extra effort and training to administer are licensed, the operational feasibility of mass vaccination campaigns for developing countries would be very limited. We applied WHO recommended routine vaccination schedules and product characteristics [17, 37], however, as of 2017, some of the routine vaccines we assumed for the simulated country had limited adoption in African Region countries (Supplemental Table 1). If we overestimated routine vaccine doses and volumes, then there would be even more available capacity for SARS-CoV-2 vaccines. Our analysis assumes 90% coverage; however, the operational needs will be decreased if there is less than 90% demand for SARS-CoV-2 vaccines among risk groups or if target coverage is lower. It is critical that pandemic vaccine response have health communications and social mobilization plans in place, and that efforts to understand and overcome vaccine hesitancy be made. While we used WHO-recommended chronic diseases prevalence estimates as risk factors for SARS-CoV-2 [14], some of these diseases may go underdiagnosed in the African Region, resulting in overestimates of vaccine demand for this risk group. We received vaccine cold storage capacity for Gavi-eligible countries. As Gavi has led the effort to strengthen cold storage capacities among countries receiving Gavi support, we do not know whether non-Gavi-eligible countries have comparable capacities to the Gavi-eligible countries which have received this support.

The development of candidate SARS-CoV-2 vaccines must take into account operational realities and programmatic suitability in developing countries. While we believe that African Region countries currently have sufficient overall cold chain capacity to accommodate SARS-CoV-2 mass vaccination campaigns, our study highlights the limitations in cold storage capacity at national levels and in workforce availability to administer these vaccines. Vaccination of risk groups with SARS-CoV-2 vaccines is possible with sufficient planning and infrastructure strengthening, but we anticipate risks to routine immunization and significant logistical challenges. Further, the development of products that are not programmatically suitable for use in developing country contexts will result in massive inequities, and concerted global efforts are urgently required to ensure that appropriate preventive interventions are made available to all who need them globally. Resources must be mobilized urgently to ensure that infrastructures and training are in place to maximize the impact of future SARS-CoV-2 vaccines in the African Region and other developing country settings.

## Data Availability

Our analysis can be replicated fully with methods and data from the study manuscript and supplement and other publicly available data which we have cited. No individual participant data were used. Other study data or documents are not available.

## ACKNOWLEDGMENTS

The work of two authors (JR and JSH) was supported by the Bill & Melinda Gates Foundation (OPP1177124). The views expressed herein are solely those of the authors and do not necessarily reflect the views of the Foundation. The work of one author (AJD) was supported by National Institute of Allergy and Infectious Diseases at National Institutes of Health (T32AI007524).

