## Supplement for "The operational impact of deploying SARS-CoV-2 vaccines in countries of the WHO African Region"

**AUTHORS AND AFFILIATIONS:**

1. Justin R. Ortiz MD*, Center for Vaccine Development and Global Health, 685 W. Baltimore St., University of Maryland School of Medicine, Baltimore, Maryland, USA.
2. Joanie Robertson*, PATH, 2201 Westlake Avenue, Suite 200, Seattle, WA 98121, USA.
3. Jui-Shan Hsu MS, PATH, 2201 Westlake Avenue, Suite 200, Seattle, WA 98121, USA.
4. Stephen L. Yu MD, Center for Vaccine Development and Global Health, 685 W. Baltimore St., University of Maryland School of Medicine, Baltimore, Maryland, USA.
5. Amanda J. Driscoll PhD, Center for Vaccine Development and Global Health, 685 W. Baltimore St., University of Maryland School of Medicine, Baltimore, Maryland, USA.
6. Sarah R. Williams MD, Division of Pulmonary and Critical Care Medicine, 110 S. Paca St, University of Maryland School of Medicine, Baltimore, Maryland, USA.
7. Wilbur H. Chen MD, Center for Vaccine Development and Global Health, 685 W. Baltimore St., University of Maryland School of Medicine, Baltimore, Maryland, USA.
8. Meagan C. Fitzpatrick PhD, Center for Vaccine Development and Global Health, 685 W. Baltimore St., University of Maryland School of Medicine, Baltimore, Maryland, USA.
9. Samba Sow, MD, Centre pour le Développement des Vaccins, Ministère de la Santé, BP251Bamako, Mali.
10. Robin J. Biellik DrPH, Independent Consultant, Tranchepied 10, 1278 La Rippe, Switzerland.
11. Jean-Marie Okwo-Bele MD, Independent Consultant, Chemin de la Poste 6, 1295 Founex, Switzerland.
12. Kathleen M. Neuzil MD, Center for Vaccine Development and Global Health, 685 W. Baltimore St., University of Maryland School of Medicine, Baltimore, Maryland, USA.

*Authors contributed equally.

Supplemental Table 1. African Region selected routine immunization programs as of 2017

| **Country^b^** | **MCV2** | **PCV** | **Rota** | **HPV** | **Rubella** | **Tetanus age 2** | **Tetanus adolescent** | **MenA** | **YF** | **BCG** |
| --- | --- | --- | --- | --- | --- | --- | --- | --- | --- | --- |
| 1. Algeria | Y | Y | N | N | Y | N | Y | N | N | Y |
| 2. Angola | Y | Y | Y | N | N | N | N | N | Y | Y |
| 3. Benin | N | Y | N | N | N | N | N | N | Y | Y |
| 4. Botswana | Y | Y | Y | Y | Y | Y | N | N | N | Y |
| 5. Burkina Faso | Y | Y | Y | N | Y | N | N | Y | Y | Y |
| 6. Burundi | Y | Y | Y | N | Y | Y | N | N | N | Y |
| 7. Cabo Verde | Y | N | N | N | Y | N | N | N | N | Y |
| 8. Cameroon | N | Y | Y | N | Y | N | N | N | Y | Y |
| 9. Central African Republic | N | Y | N | N | N | N | N | Y | Y | Y |
| 10. Chad | N | N | N | N | N | N | N | Y | Y | Y |
| 11. Comoros | N | N | N | N | N | N | N | N | N | Y |
| 12. Congo | N | Y | Y | N | N | N | N | N | Y | Y |
| 13. Côte d'Ivoire | N | Y | Y | N | N | N | N | N | Y | Y |
| 14. Democratic Republic of the Congo | N | Y | N | N | N | N | N | N | Y | Y |
| 15. Equatorial Guinea | N | N | N | N | N | N | N | N | Y | Y |
| 16. Eritrea | Y | Y | Y | N | Y | N | N | N | N | Y |
| 17. Eswatini | Y | Y | Y | N | Y | N | N | N | N | Y |
| 18. Ethiopia | N | Y | Y | N | N | N | N | N | N | Y |
| 19. Gabon | N | N | N | N | N | N | N | N | Y | Y |
| 20. Gambia | Y | Y | Y | N | Y | Y | N | N | Y | Y |
| 21. Ghana | Y | Y | Y | N | Y | N | N | Y | Y | Y |
| 22. Guinea | N | N | N | Y | N | N | N | N | Y | Y |
| 23. Guinea-Bissau | N | Y | Y | N | N | N | N | N | Y | Y |
| 24. Kenya | Y | Y | Y | N | Y | N | N | N | N | Y |
| 25. Lesotho | Y | Y | Y | N | Y | Y | N | N | N | Y |
| 26. Liberia | N | Y | Y | N | N | N | N | N | Y | Y |
| 27. Madagascar | N | Y | Y | N | N | N | N | N | N | Y |
| 28. Malawi | Y | Y | Y | N | Y | N | N | N | N | Y |
| 29. Mali | N | Y | Y | N | N | N | N | Y | Y | Y |
| 30. Mauritania | N | Y | Y | N | N | N | N | N | N | Y |
| 31. Mauritius | Y | Y | Y | Y | Y | Y | N | N | N | N |
| 32. Mozambique | Y | Y | Y | Y | N | N | N | N | N | Y |
| 33. Namibia | Y | Y | Y | N | Y | N | Y | N | N | Y |
| 34. Niger | Y | Y | Y | N | N | N | N | Y | Y | Y |
| 35. Nigeria | N | Y | N | N | N | N | N | N | Y | Y |
| 36. Rwanda | Y | Y | Y | Y | Y | N | N | N | N | Y |
| 37. Sao Tome and Principe | Y | Y | Y | Y | Y | N | N | N | Y | Y |
| 38. Senegal | Y | Y | Y | N | Y | N | N | N | Y | Y |
| 39. Seychelles | Y | N | Y | Y | Y | Y | N | N | N | Y |
| 40. Sierra Leone | Y | Y | Y | N | N | N | N | N | Y | Y |
| 41. South Africa | Y | Y | Y | Y | N | N | Y | N | N | Y |
| 42. South Sudan | N | N | N | N | N | N | N | N | N | Y |
| 43. Togo | N | Y | Y | N | N | N | N | N | Y | Y |
| 44. Uganda | N | Y | N | Y | N | N | N | N | N | Y |
| 45. Tanzania | Y | Y | Y | N | Y | N | N | N | N | Y |
| 46. Zambia | Y | Y | Y | N | Y | N | N | N | N | Y |
| 47. Zimbabwe | Y | Y | Y | Y | Y | Y | N | N | N | Y |
| Total | 26 | 39 | 34 | 10 | 22 | 7 | 3 | 6 | 23 | 46 |
| % | 55.3% | 83.0% | 72.3% | 21.3% | 46.8% | 14.9% | 6.4% | 12.7% | 48.9% | 97.9% |

Notes:

a. Some more common routine vaccines were excluded from this table, including diphtheria-tetanus-pertussis, hepatitis B, and *Haemophilus influenzae* type b vaccines.
b. Abbreviations: MCV2=second dose of measles-containing vaccine, PCV=pneumococcal conjugate vaccine, Rota=rotavirus vaccine, HPV=human papillomavirus vaccine, MenA= Neisseria meningitidis group A vaccine, YF= yellow fever vaccine, BCG= bacille Calmette-Guérin vaccine
c. Data are from 47 African Region countries reporting 2017 data to the WHO UNICEF Joint Reporting Form (JRF) on Immunization [1].

Supplemental Table 2. Vaccine doses and doses per vaccinator for routine and SARS-CoV-2 vaccination programs by nurse density estimate for WHO Regions

| **Source of nurse density input** | **Nurse density per 10,000 population** | **Routine doses / vaccinator(baseline)** | **Routine doses / vaccinator (during pandemic)** | **≥65 years (SARS-CoV-2)** | | **Chronic diseases (SARS-CoV-2)** | | **HCWs (SARS-CoV-2)** | | **All risk groups combined (SARS-CoV-2)** | | **Comparative workload between regions** |
| --- | --- | --- | --- | --- | --- | --- | --- | --- | --- | --- | --- | --- |
|  |  |  |  | **Doses / vaccinator** | **% of baseline routine for African Region** | **Doses / vaccinator** | **% of baseline routine for African Region** | **Doses / vaccinator** | **% of baseline routine for African Region** | **Doses / vaccinator** | **% of baseline routine for African Region** | **% WHO Regional Office divided by simulated country SARS-CoV-2 vaccine doses / vaccinator** |
| African Region Country | 6.9 | 165.3 | 179.7 | 48.5 | 29.3% | 164.7 | 99.6% | 2.0 | 1.2% | 186.3 | 112.7% | -- |
| Comparison using nurse density from other WHO Regions |  |  |  |  |  |  |  |  |  |  |  |  |
| Americas | 25.7 | 44.5 | 48.4 | 13.0 | 7.9% | 44.4 | 26.8% | 0.5 | 0.3% | 50.2 | 30.3% | 26.9% |
| Eastern Mediterranean | 18.9 | 60.5 | 65.8 | 17.7 | 10.7% | 60.3 | 36.5% | 0.7 | 0.4% | 68.2 | 41.3% | 36.6% |
| Europe | 66.5 | 17.2 | 18.7 | 5.0 | 3.1% | 17.1 | 10.4% | 0.2 | 0.1% | 19.4 | 11.7% | 10.4% |
| South-East Asia | 16.4 | 69.8 | 75.8 | 20.4 | 12.4% | 69.5 | 42.0% | 0.9 | 0.5% | 78.6 | 47.6% | 42.2% |
| Western Pacific | 34.9 | 32.8 | 35.6 | 9.6 | 5.8% | 32.7 | 19.8% | 0.4 | 0.2% | 36.9 | 22.3% | 19.8% |
| Global | 28.5 | 40.1 | 43.6 | 11.8 | 7.1% | 40.0 | 24.2% | 0.5 | 0.3% | 45.2 | 27.4% | 24.3% |

Notes:

a. Nurse density data are from WHO Global Health Workforce Statistics. We used the median value from countries per WHO Region at the most recent available date and divided by country population estimates to calculate per capita values [2, 3].
b. Total vaccinators is the nurse density multiplied by the simulated country population (20,000,000) x 46% (the estimated percentage of nurses that provide immunization services) [3, 4].
c. Adjustments account for 3% absenteeism (baseline) and an additional 8% absenteeism (during pandemic) [5, 6].
d. The relationship of SARS-CoV-2 vaccine workload between the estimates for the simulated country using African Region versus other WHO Regional Office nurse density estimates is the same regardless of target group.

**Supplemental Table 3. Vaccine doses and doses per vaccinator for routine and SARS-CoV-2 vaccination programs by nurse density estimate for WHO Regions**

| **African Region nurse density** | **Nurse density per 10,000 population^a^** | **Routine doses / vaccinator^c^ (baseline)** | **Routine doses / vaccinator (during pandemic)** | **≥65 years (SARS-CoV-2)** | | **Chronic diseases (SARS-CoV-2)** | | **HCWs (SARS-CoV-2)** | | **All risk groups combined (SARS-CoV-2)** | |
| --- | --- | --- | --- | --- | --- | --- | --- | --- | --- | --- | --- |
|  |  |  |  | **Doses / vaccinator** | **% of baseline routine for African Region** | **Doses / vaccinator** | **% of baseline routine for African Region** | **Doses / vaccinator** | **% of baseline routine for African Region** | **Doses / vaccinator** | **% of baseline routine for African Region** |
| 25th percentile | 4.5 | 252.0 | 273.9 | 73.9 | 29.3% | 251.1 | 99.6% | 3.1 | 1.2% | 284.0 | 112.7% |
| 50th percentile | 6.9 | 165.3 | 179.7 | 48.5 | 29.3% | 164.7 | 99.6% | 2.0 | 1.2% | 186.3 | 112.7% |
| 75th percentile | 13.8 | 83.2 | 90.4 | 24.4 | 29.3% | 82.9 | 99.6% | 1.0 | 1.2% | 93.7 | 112.7% |
| 100th percentile | 68.4 | 16.7 | 18.2 | 4.9 | 29.3% | 16.7 | 99.6% | 0.2 | 1.2% | 18.8 | 112.7% |

Notes:

a. Nurse density data are from WHO Global Health Workforce Statistics. We used the quartile and upper range value from countries in the WHO African Region at the most recent available date and divided by country population estimates to calculate per capita values [2, 3].
b. Total vaccinators is the nurse density multiplied by the simulated country population (20,000,000) x 46% (the estimated percentage of nurses that provide immunization services) [3, 4].
c. Adjustments account for 3% absenteeism (baseline) and an additional 8% absenteeism (during pandemic) [5, 6].

Supplemental Figure 1. Schematic describing vaccine flow-down through the immunization system

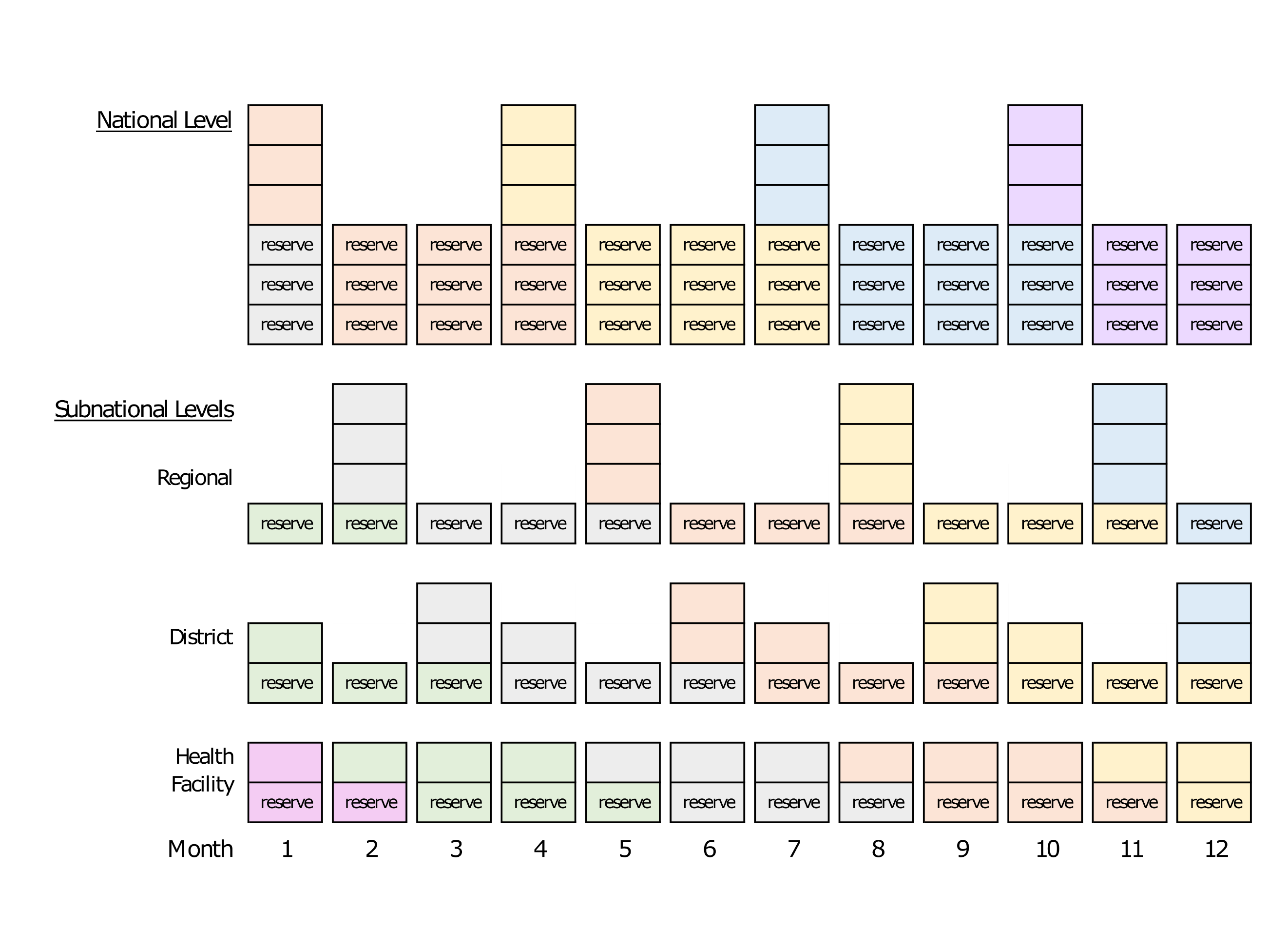

Notes:

1. Each color represents a supply shipment as it moves through the immunization system.
2. Each box represents routine vaccine doses to immunize the target population in one month, except reserve stock boxes at the health facility level which represent routine vaccine doses for 0.5 months.
3. Resupply occurs every three months at national, regional, and district levels, and monthly at the health facility level.
4. Reserve stock (excess supply in case of increased demand or stock outs) is three months at the national level, one month at regional and district levels, and 0.5 months at health facility level.
5. Vaccines stored at the national level use tertiary packaging volumes while vaccines stored at subnational levels use secondary packaging volumes. This schematic does not depict stock rotations which would preferentially use vaccines received earlier.

Supplemental Figure 2. National level monthly volumes stored for routine and SARS-CoV-2 risk group vaccination

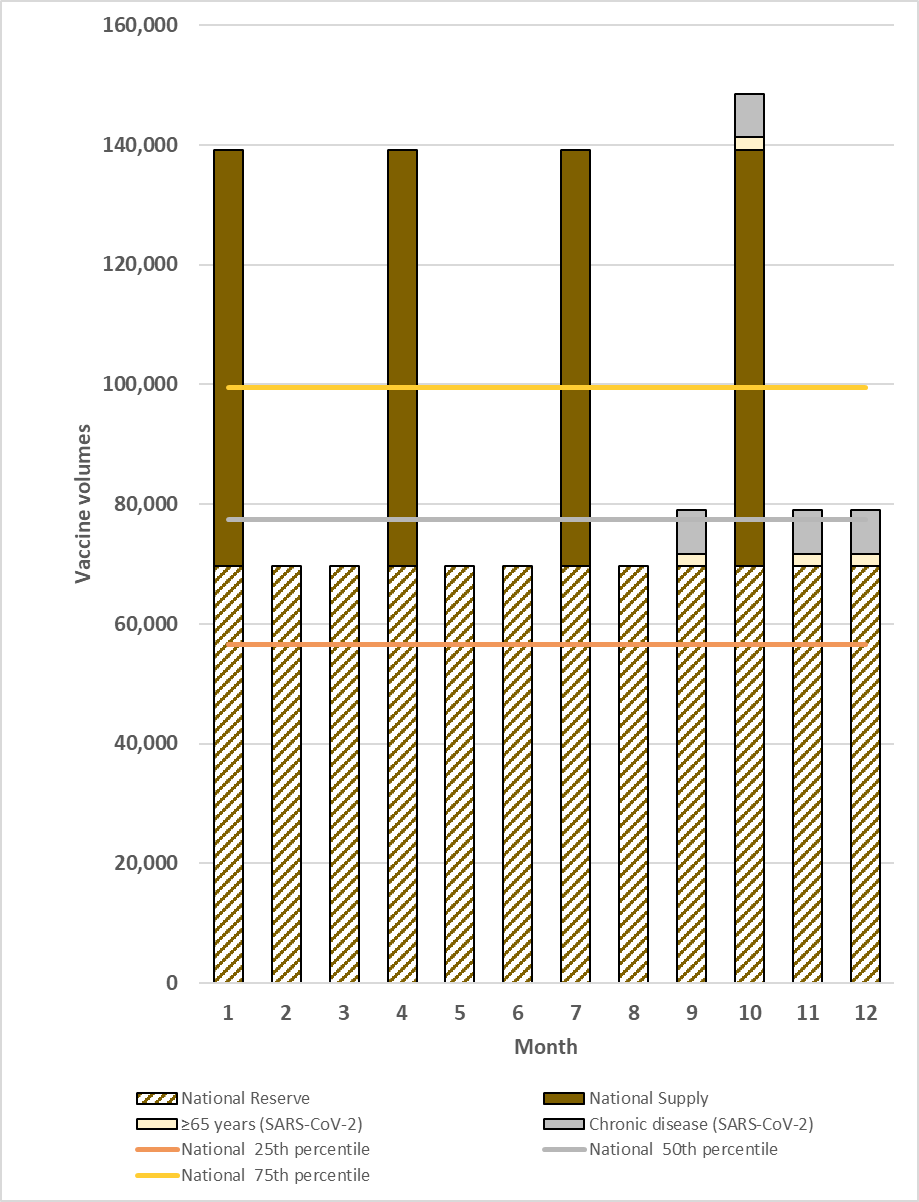

Notes:

1. Figure depicts total national level vaccine volumes calculated according to study assumptions and the vaccine flow down schematic (Figure 1).
2. The highest volume month (month 10) was chosen for the maximum national level monthly analyses.
3. HCWs target group removed because volume contribution was too small to depict in the figure.
4. Lower and upper range capacity excluded from this figure

Supplemental Figure 3. Subnational level monthly volumes stored for routine and SARS-CoV-2 risk group vaccination

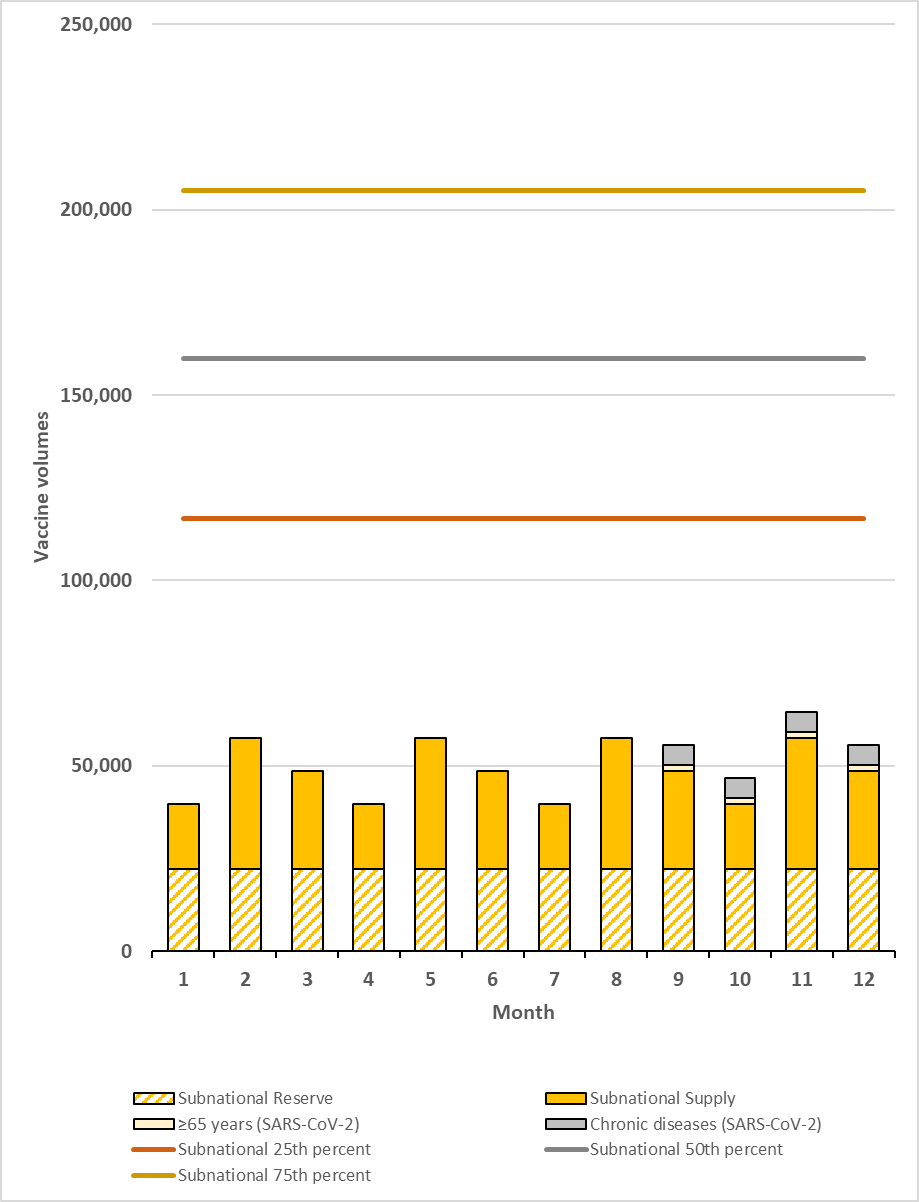

Notes:

1. Figure depicts total national level vaccine volumes calculated according to study assumptions and the vaccine flow down schematic (Figure 1).
2. The highest volume month (month 11) was chosen for the maximum national level monthly analyses.
3. HCWs target group removed because volume contribution was too small to depict in the figure.
4. Lower and upper range capacity excluded from this figure

**Supplemental Figure 4. Monthly routine and SARS-CoV-2 campaign vaccine doses per vaccinator, by quartile and upper range African Region country nurse density**

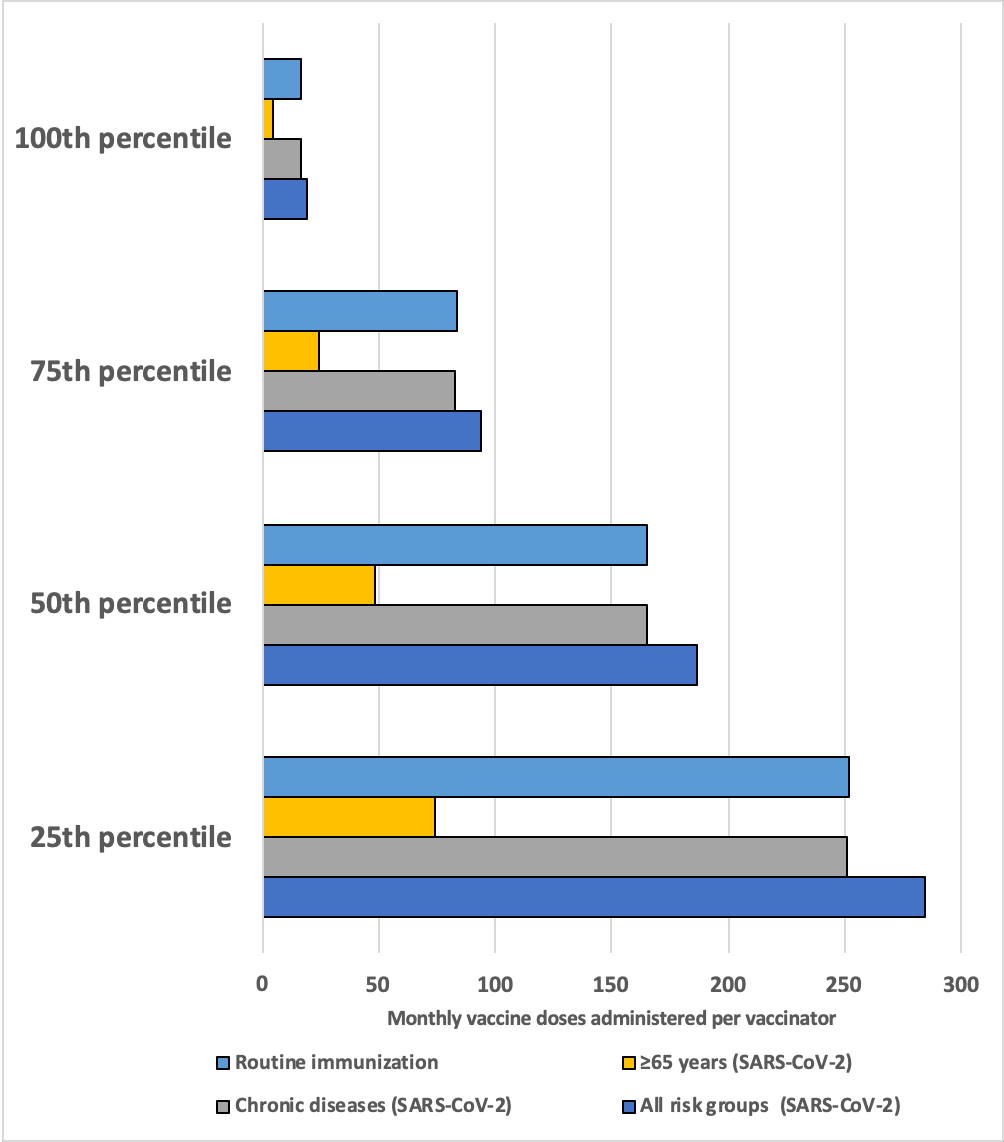

Notes:

a. Data points represent the total monthly vaccines delivered divided by the total number of vaccinators. Vaccinators = nurse density per capita x country population (20,000,000) x %nurses that provide immunization services (46%).
b. Nurse data are from WHO Global Health Workforce Statistics [3]. We used the value from countries at the most recent available date and divided by 2017 country population estimates[2] to calculate per capita values. Not all countries were represented in the WHO dataset.
c. Routine immunization programs account for 3% absenteeism (baseline). SARS-CoV-2 vaccination programs account for 3% absenteeism (baseline) and an additional 8% absenteeism (during pandemic).
d. HCWs target group removed because volume contribution was too small to depict in the figure.
